# On the Distribution of Expected Case Reproductive Number in Compartmental Disease Models

**DOI:** 10.64898/2026.11.20.26368602

**Authors:** Azadeh Aghaeeyan, Tapan Goel, Michael Roswell, Joshua S. Weitz, Jonathan Dushoff

**Affiliations:** Department of Biology, McMaster University, Hamilton, Ontario, Canada; Department of Biology, University of Maryland, College Park, Maryland, USA; University of Maryland Institute for Health Computing, North Bethesda, Maryland, USA

## Abstract

Heterogeneity in the expected number of secondary cases caused by infectors, *ℛ*_c_, has long been characterized using a skewed distribution with mean *ℛ*_0_, typically under the assumption of a fully susceptible population. We have recently shown that over the entire course of an SIR-modeled outbreak, the variation in *ℛ*_c_ is constant and independent of *ℛ*_0_. Here, we extend that result by showing that the distribution of *ℛ*_c_ across infectors over the course of a canonical SIR epidemic is precisely the standard exponential distribution, corresponding to a mean-one geometric distribution of realized infections in the canonical stochastic version. This result is exact in the limit of a small initial infection, and also requires a long-time limit in the case of persistent outbreaks. This result also applies to extensions of the model that allow immune waning and changes in *ℛ*_0_ across time (or as a function of susceptible density), but only if the infectious period remains exponentially distributed. The distribution of *ℛ*_c_, varies with *ℛ*_0_ when the generation-interval distribution is not exponential. Our findings highlight the critical role of the infectious period distribution in shaping the long-term outcomes of an outbreak.

## 1 Introduction

The basic reproduction number *ℛ*_0_, defined as the mean number of secondary infections generated by a typical infectious individual in a fully susceptible population, is a primary metric used to characterize outbreaks. However, there is substantial heterogeneity in the number of secondary infections per infector around *ℛ*_0_, even under similar environmental conditions [15, 16]. Superspreading events, in which a small fraction of infectors are responsible for a disproportionate number of infections, exemplify this heterogeneity and highlight the limitations of using a single value to describe secondary case numbers [11, 13].

Additional approaches to characterize transmission heterogeneity have been introduced to address the limitations of relying solely on *ℛ*_0_ to characterize outbreaks. The principal approach is to consider a distribution for the number of secondary cases *ℛ*_c_, referred to as the *secondary-case distribution* or *offspring distribution*. The secondary-case distribution is often modeled as a Poisson mixture, where the mixing distribution represents the variability in *individual reproductive numbers*, reflecting heterogeneity in infectiousness. The negative binomial distribution, which arises as a Poisson mixture with a gamma distribution, is commonly used for the secondary-case distribution because it allows the variance to exceed the mean, unlike the Poisson model [8].

Secondary-case distributions are widely used to quantify transmission heterogeneity across pathogens, assess outbreak potential, and evaluate the effectiveness of mitigation strategies [14]. As a result, considerable research has focused on estimating the dispersion parameter and evaluating how structured populations, time-varying contact rates, different waves of epidemic, pathogen phylogeny data influence its estimation [3, 5–7, 17]. Secondary-case distributions are frequently combined with branching-process models to derive extinction probabilities and containment metrics [2].

A key limitation of approaches grounded in branching-process theory is that they often neglect depletion of susceptible individuals. While the assumption of a constant susceptible pool is plausible in the early stages of an outbreak, it becomes less accurate as incidence accumulates. In contrast, compartmental SIR (Susceptible–Infectious–Recovered) models explicitly capture changes in the susceptible population. One approach to unify consideration of heterogeneity is through estimation of the distributions of the *case reproductive number*, which accounts for both disease dynamics and individual reproductive numbers, and can be defined as the expected number of secondary cases individual causes during their infectious period. We recently reported that, in simple SIR models, the mean and variance of the expected case reproductive number are invariant of *ℛ*_0_ when calculated over all individuals infected throughout an outbreak [12].

In this manuscript, we extend those results to show that indeed, the distribution of the expected case reproductive number *ℛ*_c_ itself is invariant across *ℛ*_0_ in an SIR model with an infinitesimal initial infectious cohort. The result carries over directly to SIR models that allow for phenomenological heterogeneity in susceptibility [4, 10]. To generalize the result to persistent outbreaks, we introduce a cutoff time and study the distribution of *ℛ*_c_ among individuals who recovered up to the cutoff time. We show this distribution is also *ℛ*_0_-agnostic provided the initial infectious cohort is negligible relative to the total number of individuals recovered by the cutoff time. We also explore how the moments of the expected case reproductive number change when the infectious period has a monotone hazard rate. Together, these results reveal the mechanistic basis and generality of unexpected invariance in transmission heterogeneity in disease outbreaks.

## 2 Results

We consider the following compartmental disease model:

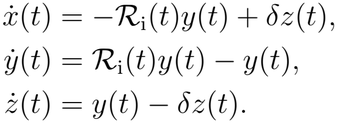

Here time is measured in the units of mean infectious period; *x* is the proportion of the population that is susceptible; *y* is the proportion of the population that is infectious; *z* is the proportion of the recovered population, *ℛ*_i_(*t*) is the instantaneous reproductive number at time *t*, and *δ* is the waning immunity rate scaled by mean infectious period.

In this model, the incidence rate, defined as the rate of new infections at time *t*, is _i_(*t*)*y*(*t*), for our autonomous (non-seasonal) model, we will use *ℛ*_i_(*t*) = *ℛ*_0_*x*^*q*+1^(*t*). The exponent *q* models the phenomenologically nonlinear dependency of incidence on the susceptible fraction. The scaled SIR model is recovered with *q* = 0 and *δ* = 0.

Individuals that become infectious at the same time *τ* and recover at the same time *ρ* constitute an *interval cohort*. The incidence weighting of such a cohort equals

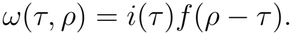

Here *f* (*·*) is the probability density function of the infectious period and equals *f* (*t*) = exp (*−t*) in the SIR model above. The expected number of infections caused by an individual infected at *τ* and recovered at *ρ*, or their expected case reproductive number, is obtained by taking the integral of the instantaneous reproductive number over the infectious period [*τ, ρ*], i.e.,

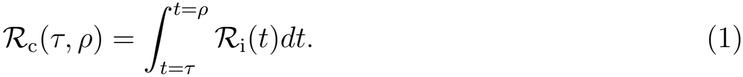

We note two points. First, *ℛ*_i_ is a population-level quantity, whereas *ℛ*_c_(*τ, ρ*) is a cohort-level quantity. Second, _c_ is an average obtained by dividing the integral by the mean infectious period, which is set to one in our model.

We are interested in the distribution of *ℛ*_c_ weighted by incidence, while ignoring intrinsic differences in transmission, seasonality, vital rates, and waning immunity (however see Discussion).

### 2.1 Distribution of *ℛ*_c_ with an exponential infectious period

To find the distribution of *ℛ*_c_, we calculate its raw moments. We denote its *k*^th^ raw moment by *ℛ*_c_^[*k*]^. The *k*^th^ raw moment of *ℛ*_c_ is defined as

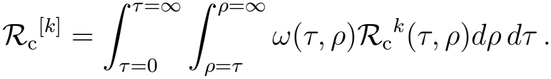

As detailed in Section S.1.1, the weighted *k*^th^ raw moment of *ℛ*_c_ is

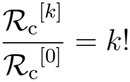

for an SIR epidemic model where the infectious period is exponentially distributed, and the initial infectious cohort is infinitesimal. This means that the distribution of the expected case reproductive number is a unit exponential [1] (Figure 1).

**Figure 1:**
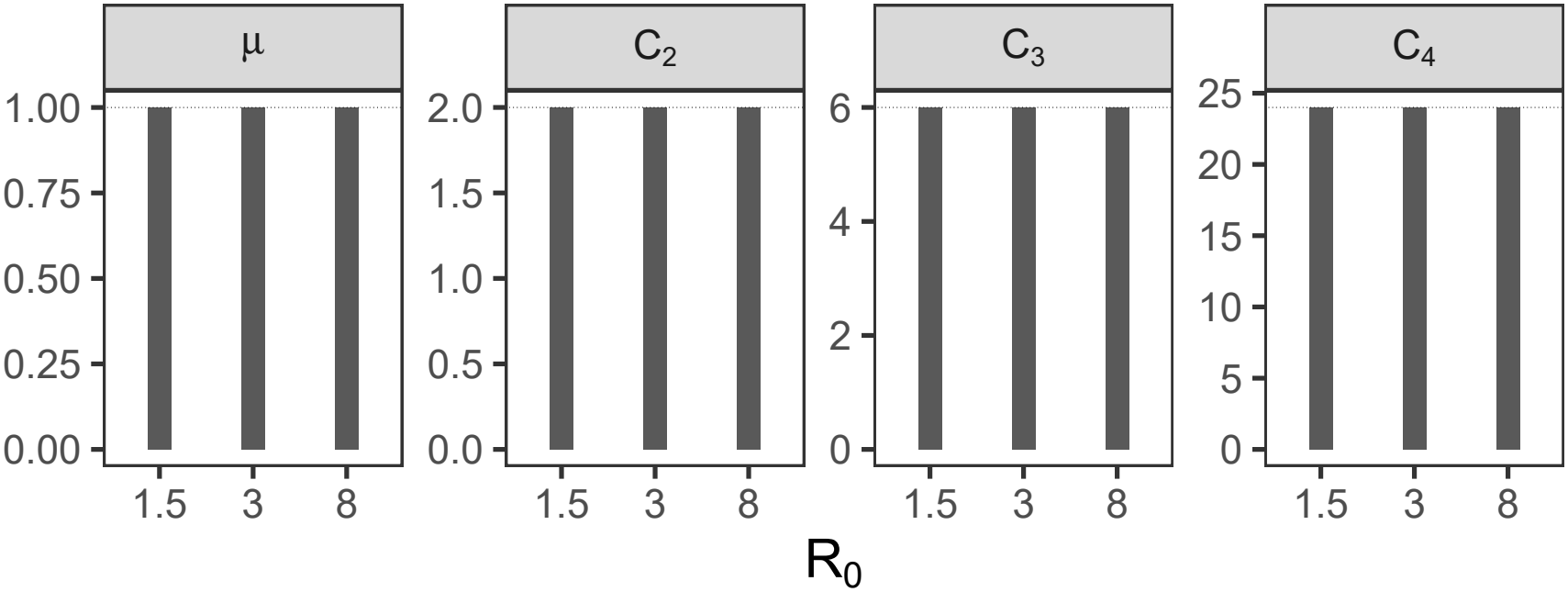
The distribution of the expected case reproductive number *ℛ*_c_ is exponential in simulations of an SIR model, regardless of *ℛ*_0_. The panels show the first four normalized raw moments of *ℛ*_c_, computed from simulations of SIR models with three different values of _0_. The horizontal line in each panel represents the corresponding raw moment for a unit exponential distribution as proved in Section S.1.1. To simulate SIR epidemics, we implemented the model described by equations S.8, with parameter values *δ* = 0, *α* = 0, *q* = 0, and *β*_0_ = 1.5, 3, 8, for 100 mean infectious periods.

In simulation studies, the weighted mean of *ℛ*_c_, denoted by *µ*, is not exactly one. We then divide the *k*^th^ weighted raw moment by the *k*^th^ power of mean to obtain the *normalized* moment:

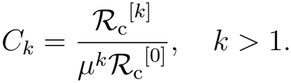

By doing so, we characterize its shape independently of scale (Figure 1).

We then introduced a cutoff time, *t*_*f*_, and showed the distribution of *ℛ*_c_ among individuals who recovered by time *t*_*f*_ is also unit exponential for sufficiently large *t*_*f*_. For a large cutoff time *t*_*f*_, the set of total infections is effectively the same as the set of recovered individuals. Thus, the results generalize to persistent outbreaks like SIRS models in a long-time limit. See Section S.1.2 for a proof.

We used the dispersion parameter *κ*, defined as the coefficient of variation squared [12], to quantify deviations of the *ℛ*_c_ distribution from an exponential distribution in the presence of a non-infinitesimal initial infectious cohort size. As shown in Figure 2, *κ* gets closer to 1 as the ratio of the initial infectious cohort size to the total number of infections approaches zero, which can occur in SIRS models. Our numerical simulations also predict a more pronounced deviation for larger values of *ℛ*_0_.

**Figure 2:**
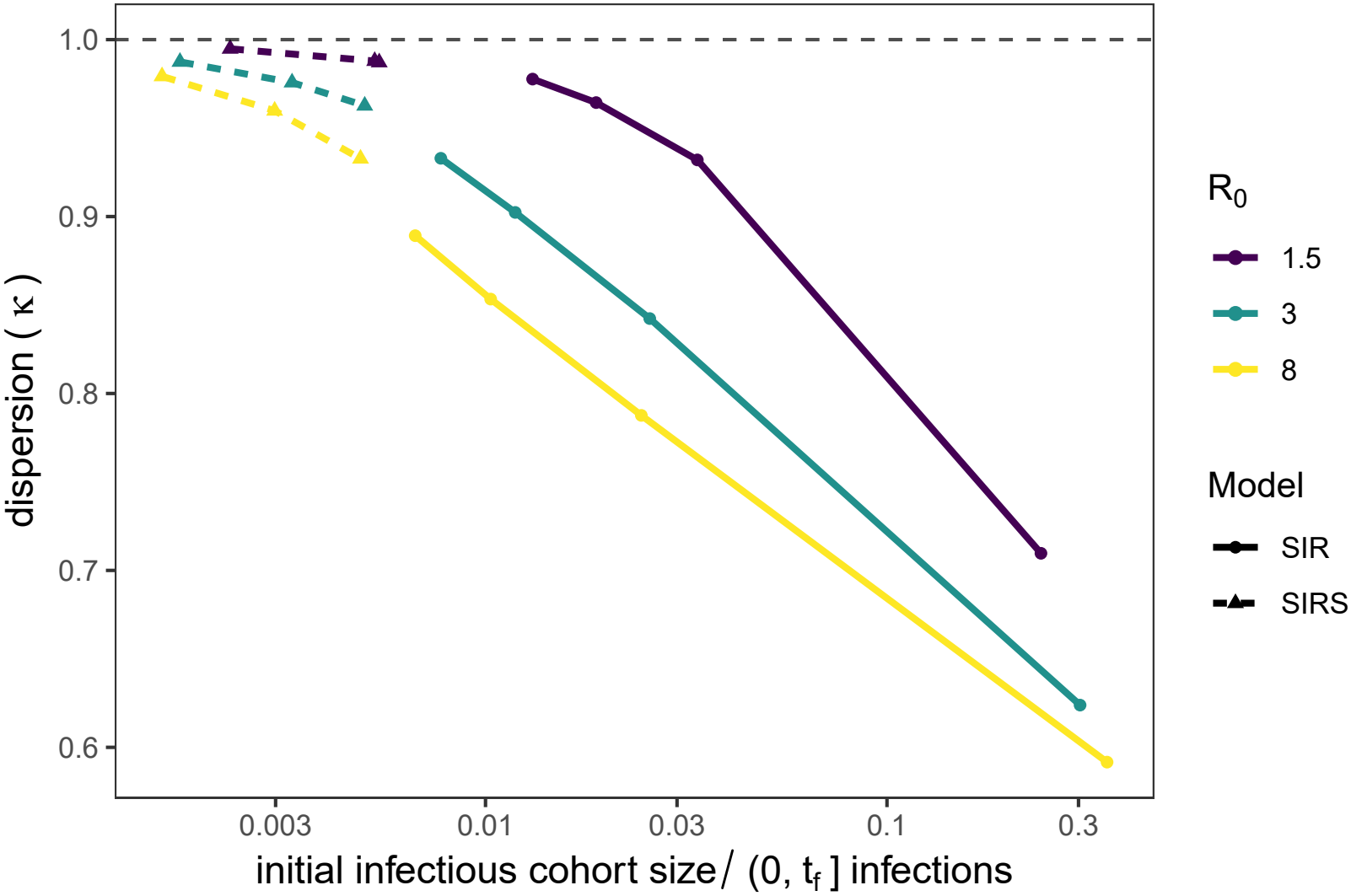
The dispersion in the expected case reproductive number with non-negligible initial infectious cohort. becomes closer to 1 as the ratio of the initial infectious cohort size to the infections occurring during (0, *t*_*f*_] approaches zero, where *t*_*f*_ is the cutoff time. This can occur in a seasonally forced SIRS epidemic, but not in a canonical SIR epidemic. We computed dispersion in *ℛ*_c_ for individuals who had recovered by time *t*_*f*_, excluding the initial infectious cohort. The seasonally forced SIRS epidemic was modeled using the equations S.8 with *δ* = 0.01, *α* = 1, *T* = 60, and *q* = 0. For each value *ℛ*_0_, we computed the dispersion for three different cutoff points *t*_*f*_ = 10, 100, and 300. We implemented the model described by equations S.8, with parameter values *δ* = 0, *α* = 0, *q* = 0 to simulate an SIR epidemic. For each value of *ℛ*_0_, the dispersion was computed for four different cutoff points *t*_*f*_ = 0.25tpeak, 0.75tpeak, tpeak, and 1.25tpeak, where tpeak is the time of peak incidence. For both models, the initial infectious proportion is *y*_0_ = 0.005, and time is measured in units of mean infectious period. The horizontal axis is on a log scale.

### 2.2 Distribution of *ℛ*_c_ with a non-exponential infectious period

Here we explored how different forms of infectious period distribution affect the *ℛ*_c_ distribution. In particular we studied infectious period distributions with increasing hazard rates and distributions with decreasing hazard rates. The Erlang distribution and the Weibull distribution with shape parameter greater than one are examples of distributions having increasing hazard rates, whereas the Weibull distribution with shape parameter less than one and the hyperexponential distribution have decreasing hazard rates.

The normalized moments of *ℛ*_c_ are bounded above by those of the unit exponential distribution when the infectious period has an increasing hazard rate (Figure 3, panel a). Conversely, when the infectious period has a decreasing hazard rate, the moments of *ℛ*_c_ are bounded below by those of the unit exponential distribution (Figure 3, panel b). Our numerical results suggest that, in both cases, when *ℛ*_0_ is larger, the moments of *ℛ*_c_ distribution are closer to those of the unit exponential distribution (Figure 3).

**Figure 3:**
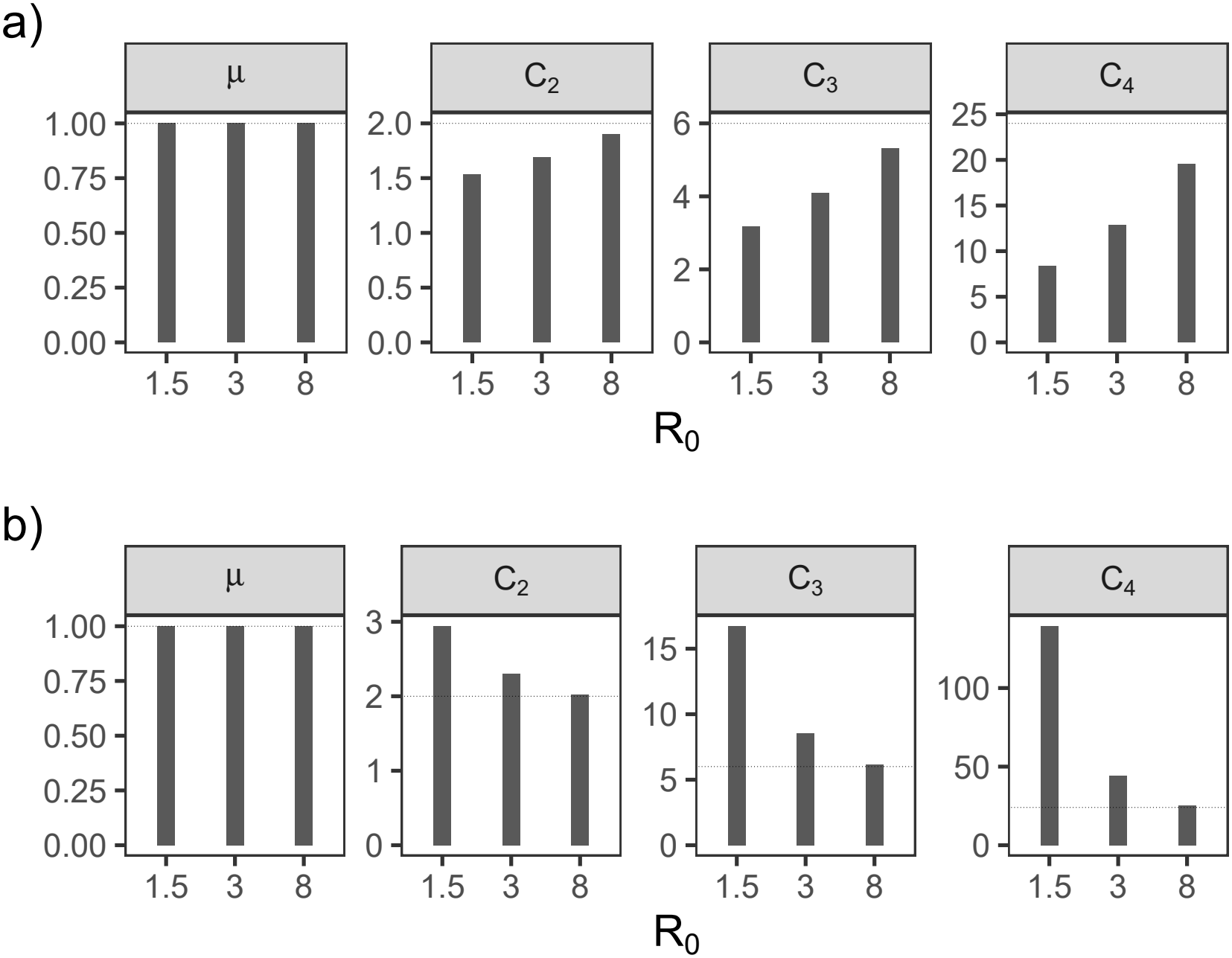
The first four normalized raw moments of *ℛ*_c_ in simulations of SIR models with infectious distributions having monotone hazard rates. Panel a shows results for an infectious period with an increasing hazard rate, and panel b for an infectious period distribution with a decreasing hazard rate. In agreement with theoretical results, for an epidemic with an infectious distribution with an increasing hazard rate, the *k*^th^ weighted raw moment of *ℛ*_c_ is bounded above by *k*!. However, in the presence of an infectious distribution with a decreasing hazard rate, the *k*^th^ weighted raw moment of expected case reproductive number is bounded below by *k*!. The horizontal line in each panel represents the corresponding moment for a unit exponential distribution. To model an infectious period with an increasing hazard rate, we used an Erlang distribution with mean one and squared coefficient of variation 1*/*2 (*m* = 2 in Equation (S.9)). To model a distribution with a decreasing hazard rate, we used a two-phase hyperexponential distribution (*γ*_1_ = 0.4, *γ*_2_ = 1.6, and *p*_1_ = 0.2 in Equation (S.10)).

Further, we proved that the presence of non-infinitesimal initial infectious cohort size reduces the *k*^th^ moment of *ℛ*_c_ (see Section S.1.1 and Section S.1.1.1 for details).

## 3 Discussion

We have shown that there is a “universal”, long-term distribution of expected cases per infector *ℛ*_c_, corresponding to a universal distribution of realized cases across infectors in simple stochastic realizations, for a broad family of SIR-based compartmental models. The expected-case distribution is the standard exponential, and the corresponding realized-case distribution is the mean-one geometric. These results hold in the limit where the number of infectors at the beginning and end is relatively small compared to the total number ever infected. For a non-recurrent epidemic this means that the number of initial infectors must be small (and typically the epidemic is taken to be complete). For a recurrent epidemic it requires only that we take a long-time limit.

The result holds for seasonal forcing in transmission, and for cases (e.g., phenomenological heterogeneity) where infection is not linear in the number of susceptibles. It also holds when immunity wanes, or when vital (birth-death) dynamics are added. It does not hold when the distribution of the generation-interval, that is, the time between when an individual becomes infected and the time they transmit the infection to another individual, is not exponential. In general, these lead to *ℛ*_c_ distributions that depend on *ℛ*_0_.

## Data Availability

All scripts used to produce the MS are available online at https://github.com/dushoff/momentsCode

## Data and code availability

All code and simulation files can be found at https://github.com/dushoff/momentsCode.

## Acknowledgments

J.S.W. is an investigator at the University of Maryland-Institute for Health Computing, which is supported by funding from Montgomery County, Maryland and The University of Maryland Strategic Partnership: MPowering the State, a formal collaboration between the University of Maryland, College Park and the University of Maryland, Baltimore. J.S.W acknowledges support from Simons Foundation grant MPS-SIP-00930382. JD and AA were supported by the Canadian Institutes for Health Research.

## S.1 Proofs

### S.1.1 Distribution of *ℛ*_c_ in non-persistent outbreaks

In this section, we investigate the distribution of *ℛ*_c_ weighted by the incidence over the entire course of an outbreak. Here, we study epidemics in which the initial infection cohort is infinitesimal, and the outbreak burns out, so the final size is bounded (see Section S.1.1.1 and Section S.1.2 for results with a non-infinitesimal initial infection cohort and a persistent outbreak, respectively).

In what follows, we show that the *k*^th^ (weighted) raw moment of *ℛ*_c_ in an epidemic model in which the infectious period is exponentially distributed equals *k*!.

To avoid excessive use of parentheses, we adopt the convention that in expressions of the form ∫∫*dτdρg*(*τ, ρ*), the leftmost differential corresponds to the outer integral. Hence,

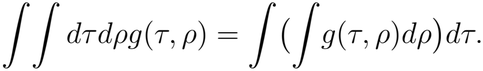

The *k*^th^ raw moment of the expected case reproductive number is

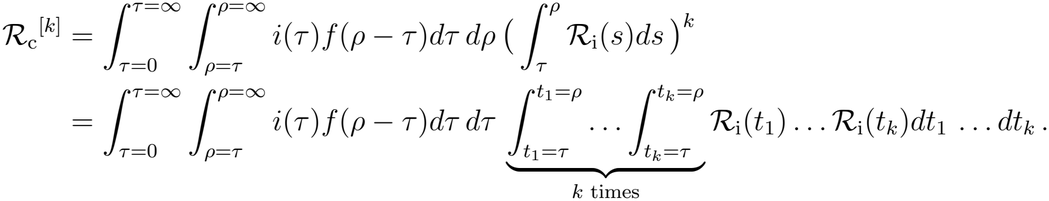

The integrand of the last *k* integrals is the symmetric *k*-variable function *ℛ*_i_(*t*_1_) … *ℛ* _i_(*t*_*k*_), and the domain of integration is the hypercube [*τ, ρ*]^*k*^. The integral is equal to *k*! times the integral of the function over the region *τ < t*_1_ *< t*_2_ *<* … *< t*_*k*_ *< ρ*. Consider any region within the hypercube where the dimensions are ordered, such as *t*_1_ *< t*_2_ *<* … *< t*_*k*_. This ordering selects *k* dimensions from the *k* available dimensions. Thus, there are *k*! possible orderings. Each possible ordering corresponds to a region of equal volume, and the sum of these volumes equals the total volume of the hypercube. Consequently, the volume of each region is 1*/k*! of the total volume:

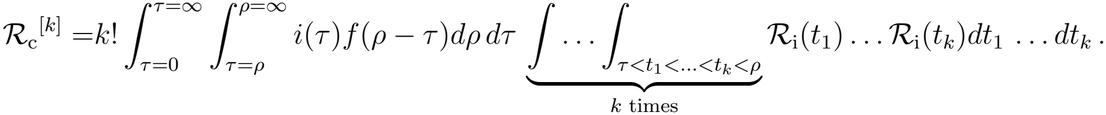

To be able to interpret the terms as epidemiological tangible quantities, we swap the integral orders. We start by taking the integral over the time of recovery *ρ*.

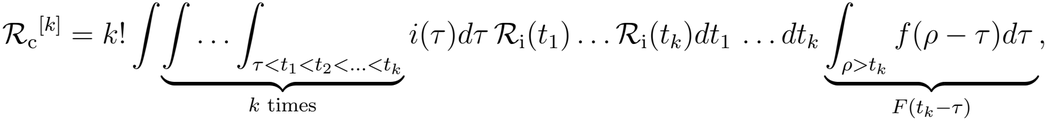

where *F* denotes the survival function associated with the density of the infectious period.

The term *F* (*t*_*k*_ *− τ*) is the proportion of the population who became infected at time *τ* and remain infectious at time *t*_*k*_. For an exponential infectious period, *F* (*t*_*k*_ *− τ*) equals the product of the proportions of individuals with infectious periods exceeding *τ −t*_1_, *−t*_2_ *t*_1_, and so forth, with the final term being the proportion of those whose infectious period exceeds *t*_*k*_ *− t*_*k−*1_. i.e.,

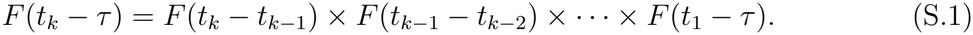

In contrast, when the infectious period has an increasing hazard rate^1^, such as the Erlang distribution, the equality (S.1) changes to the inequality, i.e.,

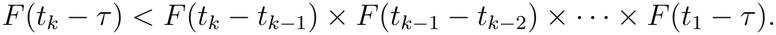

For an infectious period has a decreasing hazard rate^2^, such as the Weibull distribution with parameter shape less than one, the equality (S.1) changes to the inequality, i.e.,

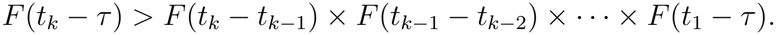

Every element in the right-hand side is non-negative, and hence

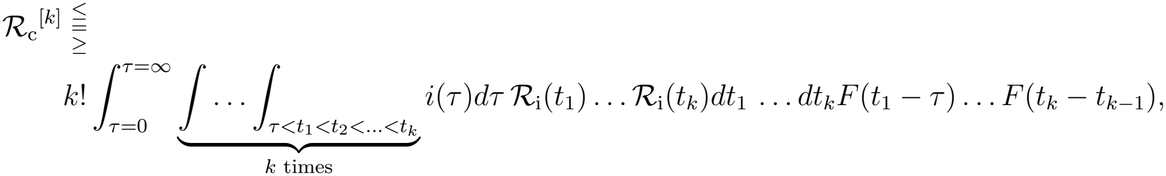

where the less-than-or-equal-to sign is valid for an infectious period with an increasing hazard rate, the equal-to sign holds for an exponentially distributed infectious period, and the greater-than-or-equal-to sign is valid for an infectious period with a decreasing hazard rate. Moving the *τ −* integration to the innermost position and *t*_*k*_ to the outermost position results in

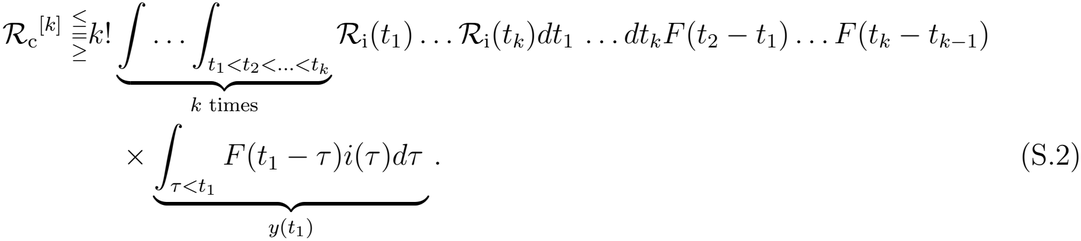

The innermost integral over *τ* adds up the weight of individuals infected at any time before *t*_1_ and still infectious at *t*_1_, which is the proportion that is infectious at *t*_1_, *y*(*t*_1_). By definition, the product of *ℛ* (*t*_*1*_) and *y*(*t*_*1*_) is *i*(*t*_*1*_). The now innermost integral, 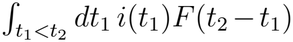, quals *y*(*t*_2_). More generally, the term 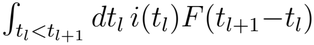, for *l ∈ {*2, …, *k−*1*}*, is equal to *y*(*t*_*l*+1_). The multiplication of *y*(*t*_*l*+1_) and *ℛ*_i_(*t*_*l*+1_) equals *i*(*t*_*l*+1_). By repeating the procedure and integrating out the variables *t*_3_, …, *t* _*k−*1_, the remaining term is 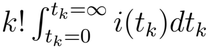.

The integral is simply *R*_c_^[0]^ and equals final size

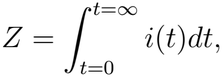

and hence

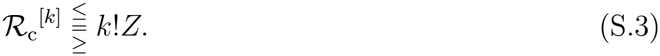

The *k*^th^ weighted raw moment is defined as

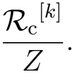

Thus, the *k*^th^ weighted raw moment is equal to *k*! for an exponentially distributed infectious period; it is bounded by *k*! for an infectious period having an increasing hazard rate, and it is lower bounded by *k*! for an infectious period having a decreasing hazard rate.

The *k*^th^ raw moment of the unit exponential distribution equals *k*! as well, and this distribution is uniquely determined by its moments [1]. This means that the distribution of the case reproductive number for an exponentially distributed infectious period, is exponential with rate 1.

#### S.1.1.1 A non-infinitesimal initial infectious population

The derived results in the previous section hold under the assumption of an infinitesimal initial infection cohort. If this assumption does not hold, the rightmost term on the RHS of Equation (S.2), will be less than the total proportion of infectious population at time *t*_1_, and hence 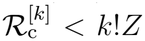. The total weight however, remains equal to *Z*. Thus, the *k*^th^ weighted raw moment of *ℛ*_c_ will be bounded above by *k*!.

### S.1.2 Distribution of *ℛ*_c_ in persistence outbreaks

The proof in Section S.1.1 hinges on the assumption that the outbreak eventually burns out and the final size is bounded. However, waning immunity can replenish the susceptible pool, allowing the outbreak to persist. To extend our the results to such settings, we introduce a cutoff time, *t*_*f*_, and study the distribution of the expected case reproductive number among individuals who became infectious and subsequently recovered during the interval (0, *t*_*f*_).

For an outbreak and a sufficiently large *t*_*f*_, the population of recovered individuals is effectively the same as the population of infected ones. We therefore expect a result similar to that in Section S.1.1.

By ℛ_c_(*t*_*f*_), we denote the excepted case reproductive number among individuals who became infected and subsequently recovered during the time interval (0, *t*_*f*_), and its *k*^th^ raw moment is given by

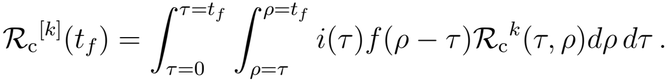

The weight of individuals who became *infected* and *recovered* during this interval reads as

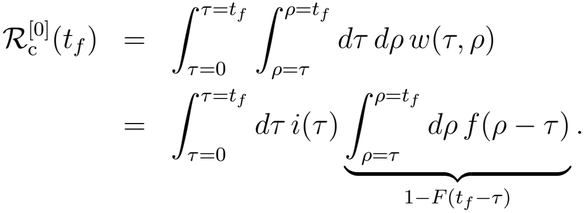

The innermost integral sums the density function for recovery period from 0 to *t*_*f*_ *− τ* which equals the proportion of infectious individuals who recovered before *t*_*f*_, i.e., 1 *− F* (*t*_*f*_ *− τ*). Plugging 1 *− F* (*t*_*f*_ *− τ*) into the right-hand side part results in

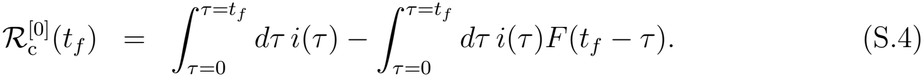

Assuming the proportion of initially infectious individuals is negligible, the first term on the right hand side equals the total proportion of individuals infected before time *t*_*f*_, denoted by *z*(*t*_*f*_), and the second term is the total proportion of infectious individuals at time *t*_*f*_, i.e., *y*(*t*_*f*_). Thus,

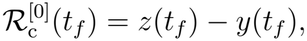

which is expected. The first moment 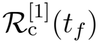 is given by

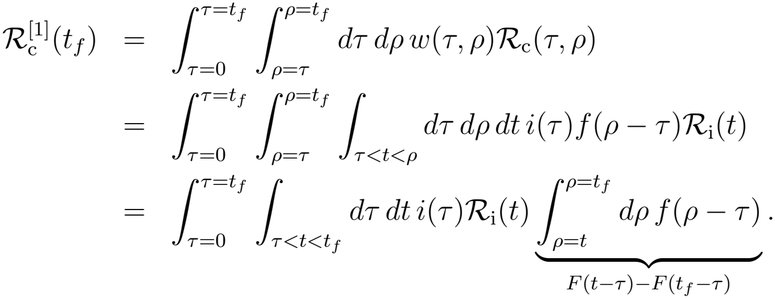

Plugging *F* (*t − τ*) *− F* (*t*_*f*_ *− τ*) into the right hand side of the equation and assuming an exponential distribution for the infectious period results in

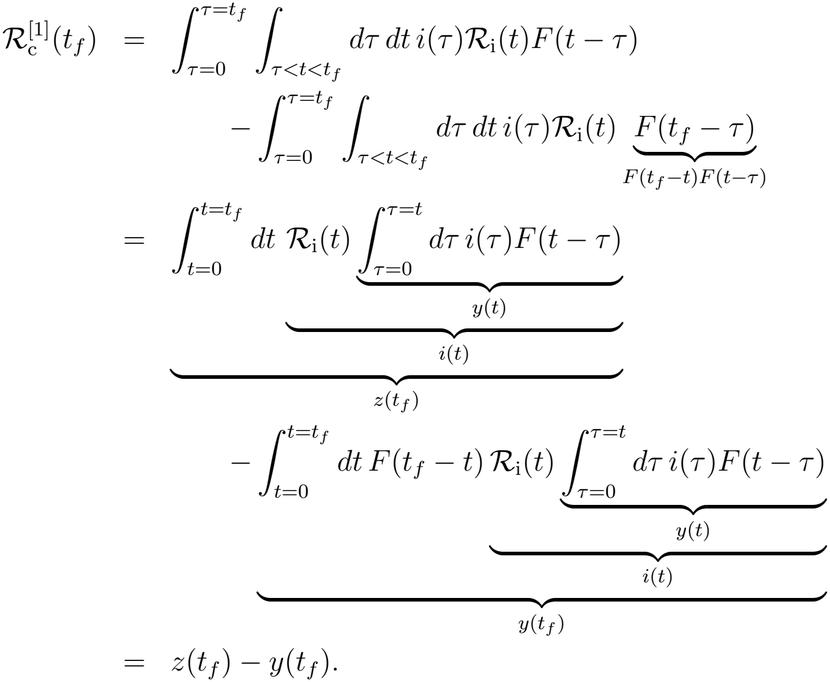

Thus the weighted first moment is equal to

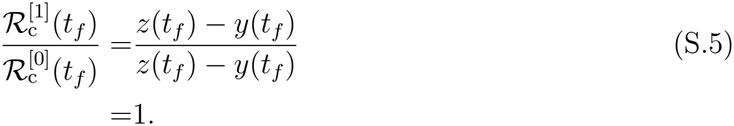

Note that the equation (S.5) is valid in the limit as the ratio of the initial infectious cohort size to the recovered cohort size 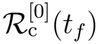 tends to zero.

The *k*^th^ raw moment of *R*_c_^[*k*]^(*t*_*f*_) is equal to

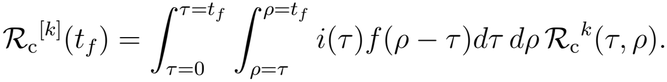

Replacing the integral over the hypercube [*τ, ρ*]^*k*^ with *k*! times the integral over the domain *t*_1_ *< t*_2_ *<* … *< t*_*f*_ and moving the *ρ−*integration to the innermost yields

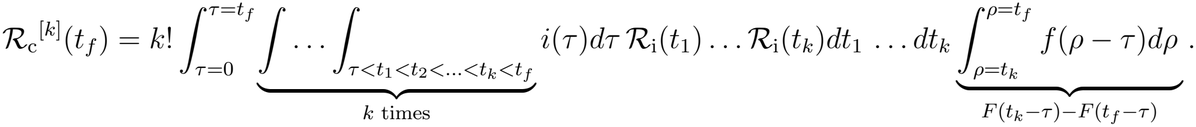

Plugging *F* (*t*_*k*_ *− τ*) *− F* (*t*_*f*_ *− τ*) into the right hand side, we have

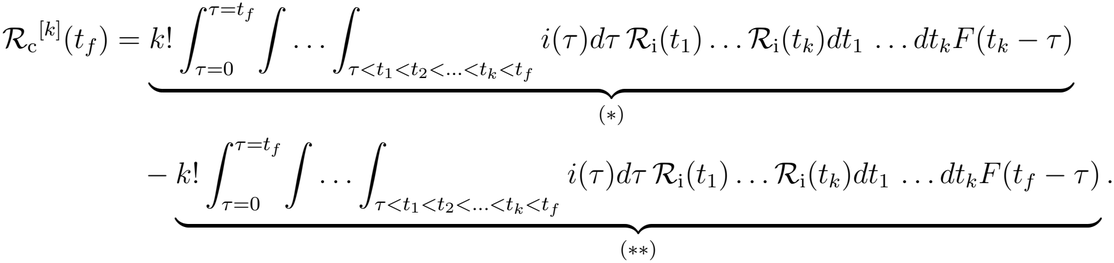

If the recovery time is exponentially distributed, then equation (S.1) holds. By substituting the right-hand side of (S.1) into (*\**), we find that (*\**) equals *k*!*z*(*t*_*f*_). As for the term (*\*\**), (S.1) takes the form

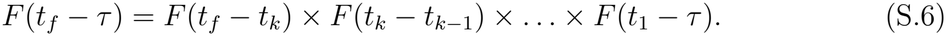

Replacing *F* (*t*_*f*_ *− τ*) with the right hand side of (S.6) in (*\*\**), it follows that (*\*\**) equals *k*!*y*(*t*_*f*_). Thus,

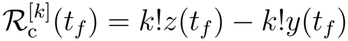

and hence the weighted *k*^th^ moment is

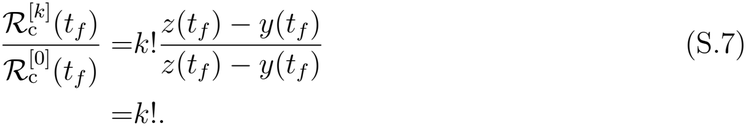

Thus, *ℛ*_c_(*t*_*f*_) is also exponentially distributed with rate one.

## S.2 Simulation Models

### SIRS Model

An SIRS compartmental model with seasonal forcing and nonlinear incidence is described as:

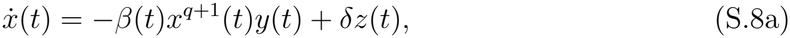

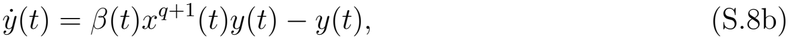

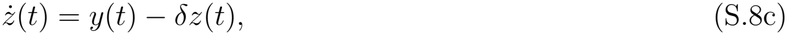

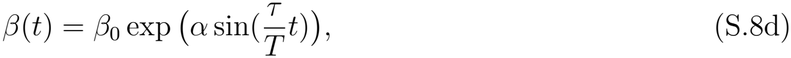

where *x* is the proportion of the population that is susceptible, *y* is the population proportion of the infected individuals, and *z* is the population proportion of recovered individuals. Time *t* is measured in units of the mean infectious period. The rate of waning immunity is denoted by *δ*. The dimensionless exponent *q* captures the phenomenologically nonlinear dependency of the incidence on the susceptible fraction. The standard mass-action term is recovered with *q* = 0. The mean transmission rate is denoted by *β*_0_, *α* is the log of the amplitude of seasonality, *τ* = 2*π*, and *T* is the seasonality period.

### SIR Model with Erlang Infectious Period

To model an Erlang-distributed infectious period, we used the linear chain method, in which the infectious compartment is divided into several sub-compartments with identical mean residence times. For an outbreak with a constant transmission rate, homogeneous susceptibility, and an infectious period with mean 1 and squared coefficient of variation 1*/m*, the model takes the form

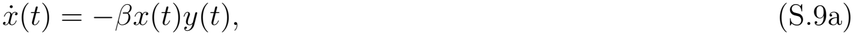

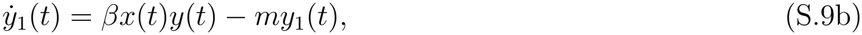

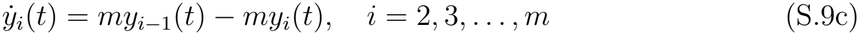

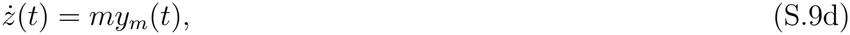

where 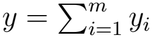.

### SIR Model with Hyper-exponential Infectious Period

An example of a distribution with a decreasing hazard rate is the hyperexponential distribution, with density

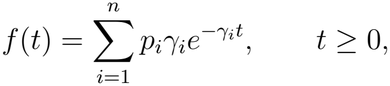

where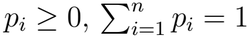, and *γ*_*i*_ *>* 0. Under this parametrization, the mean is

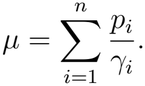

A hyperexponential distribution always has a decreasing hazard rate, except in the degenerate case *γ*_*i*_ = *γ* for *i* = 1, …, *n*, in which case it reduces to an exponential distribution.

To model an outbreak with a hyperexponentially distributed infectious period, we use the following set of ODEs:

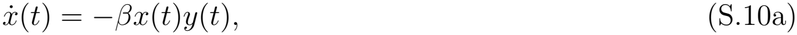

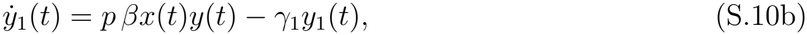

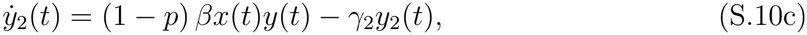

where *y* = *y*_1_ + *y*_2_.

## Footnotes

1 The hazard function for a density function *f* (*t*) with associated survival function *F* (*t*) and non-negative support equals *f* (*t*)*/F* (*t*). In an infectious distribution with an increasing hazard rate (IHR), the longer an individual has been infectious, the higher their conditional instantaneous recovery rate. The survival function associated with IHR class satisfies *F* (*a* + *b*) *< F* (*a*)*F* (*b*) [9].

2 In an infectious distribution with a decreasing hazard rate (DHR), the longer an individual has been infectious, the lower their conditional instantaneous recovery rate. The survival function associated with DHR class satisfies *F* (*a* + *b*) *> F* (*a*)*F* (*b*).

